# Prognostic value of the mitral E/e’ ratio during exercise in asymptomatic patients with low-gradient severe aortic stenosis and preserved ejection fraction

**DOI:** 10.1101/2023.11.15.23298603

**Authors:** Daisuke Miyahara, Masaki Izumo, Yukio Sato, Tatsuro Shoji, Risako Murata, Ryutaro Oda, Taishi Okuno, Shingo Kuwata, Yoshihiro J Akashi

**Author notes:** Address for Correspondence: Masaki Izumo, MD, PhD., 2-16-1, Sugao, Miyamae-ku, Kawasaki, 216-8511, Japan.

## Abstract

**Background:** Current evidence on the prognostic value of exercise stress echocardiography (ESE) in asymptomatic patients with low-gradient severe AS is limited. Therefore, this study aimed to elucidate its prognostic implications for patients with low-gradient severe AS and determine the added value of ESE in risk stratification for this population.

**Methods:** This retrospective observational study included 122 consecutive asymptomatic patients with either moderate (mean pressure gradient [MPG] <40 mmHg and aortic valve area [AVA] 1.0–1.5 cm^2^) or low-gradient severe (MPG <40 mmHg and AVA <1.0 cm^2^) AS and preserved left ventricular ejection fraction (≥50%) who underwent ESE. All patients were followed up for AS-related events.

**Results:** Of 143 patients, 21 who met any exclusion criteria, including early interventions, were excluded, and 122 conservatively managed patients (76.5 [71.0–80.3] years; 48.3% male) were included in this study. During a median follow-up period of 989 (578–1571) days, 64 patients experienced AS-related events. Patients with low-gradient severe AS had significantly lower event-free survival rates than those with moderate AS (log-rank test, p<0.001). Multivariable Cox regression analysis showed that the mitral E/e’ ratio during exercise was independently associated with AS-related events (hazard ratio=1.075, p<0.001) in patients with low-gradient severe AS.

**Conclusions:** This study suggests that asymptomatic patients with low-gradient severe AS have worse prognoses than those with moderate AS. Additionally, the mitral E/e’ ratio during exercise is a useful parameter for risk stratification in patients with low-gradient severe AS.

**Clinical Perspective:** Aortic stenosis (AS) is increasingly prevalent in aging society, and risk stratification of patients with low-gradient severe AS and preserved ejection fraction remains controversial. Exercise testing has been useful for identifying symptoms or abnormal hemodynamic responses during exercise in patients with asymptomatic AS. Limited evidence supports exercise stress echocardiography (ESE) as a valuable tool for evaluating asymptomatic AS. This study investigated the prognoses of patients with low-gradient severe AS and the utilization of ESE for risk stratification of these patients. Patients with low-gradient severe AS had worse prognoses than those with moderate AS. The mitral E/e’ ratio during exercise can be utilized as a parameter for risk stratification of patients with low-gradient severe AS.

## Introduction

Aortic stenosis (AS) is a prevalent valvular heart disease, particularly in developing countries, and its incidence increases with an aging population (1,2), affecting 2–9% of older persons (2–4). Echocardiography serves as the cornerstone of evaluating and grading AS. AS severity is generally determined based on aortic valve area (AVA) and mean pressure gradient (MPG). Discordant echocardiographic measurements are occasionally encountered; some patients may have severe stenosis based on the AVA but not severe stenosis based on MPG, even when left ventricular ejection fraction (LVEF) is preserved (5).

Previous studies have indicated that patients with low-flow, low-gradient, severe AS represent a subgroup with advanced-stage AS, reduced stroke volume (SV), and poor prognoses (6–9). Conversely, another study indicated that the prognoses of these patients were similar to those of patients with moderate AS (10). Symptomatic patients with paradoxical low-flow low-gradient (PLFLG) severe AS have been reported to have poor prognoses in many studies, and guidelines recommend invasive therapy for stage D valvular heart disease (11). However, this inconsistency between MPG and AVA is common in daily clinical practice, making proper classification of AS severity challenging. Therefore, the prognostic implications of low-gradient severe AS remain controversial and are not fully understood, and most previous studies have relied on echocardiographic measurements performed at rest.

Exercise testing has demonstrated the utility of identifying symptoms or abnormal hemodynamic responses during exercise in patients with asymptomatic AS. Exercise stress echocardiography (ESE) provides additional information on the flow and pressure dynamics associated with AS. However, limited evidence supports ESE as a valuable tool for evaluating asymptomatic AS. Previous studies have reported that increased MPG levels during exercise and exercise-induced pulmonary hypertension (EIPH) were important indicators of symptom development and adverse outcomes. However, other studies have questioned the efficacy of these findings in risk stratification, possibly because they are influenced by contractile reserves and myocardial responses. Therefore, this study aimed to elucidate the prognoses of asymptomatic patients with low-gradient severe AS in comparison with those with moderate AS and explore additional information ESE can offer for risk stratification in this cohort.

## Methods

### Study design

This study retrospectively observed 143 consecutive asymptomatic patients with either moderate (MPG <40 mmHg and AVA 1.0–1.5 cm^2^) or low-gradient severe (MPG <40 mmHg and AVA <1.0 cm^2^) AS and preserved LVEF (≥50%) who underwent ESE between January 2013 and December 2021 at St. Marianna University Hospital. All patients in this study are at least 20 years old. The study protocol was approved by the ethics committee of St. Marianna University School of Medicine (No. 6250), and the need for informed consent was waived due to the retrospective nature of the study.

### Resting echocardiography

All patients underwent comprehensive two-dimensional and Doppler transthoracic echocardiography before exercise testing in accordance with the American Society of Echocardiography guidelines (12). An experienced sonographer performed all echocardiographic procedures using Vivid E9 or E95 ultrasound systems (General Electric Healthcare, Little Chalfont, UK). The left ventricular (LV) end-diastolic volume (EDV) and end-systolic volume (ESV) were measured using the Simpson biplane method. The LVEF was calculated as follows: [(EDV-ESV)/EDV] × 100. Continuous-wave Doppler was used to measure the maximal aortic valve velocities in apical three or five chamber views, and the peak and mean gradients were estimated based on the Simplified Bernoulli equation. The LV outflow tract (LVOT) diameter was measured using zoomed parasternal long-axis views. The LVOT velocities acquired using pulsed-wave Doppler and the velocity-time integrals (VTI LVOT) were measured.

The SV was calculated using the following formula: (LVOT diameter/2)^2^ × 3.14 × VTI LVOT. Subsequently, the SV index (SVi) and cardiac output (CO) were estimated using the following formulas: SVi=SV/body surface area (BSA) and CO=SV × heart rate (HR). The AVA was calculated using the continuity equation, and the AVA index was calculated by dividing it with the BSA. The global LV afterload was estimated using the valvulo-arterial impedance (Zva), formulated as follows: Zva=(systolic blood pressure + MPG)/SVi (13). The systolic pulmonary artery pressure (SPAP) was derived from the regurgitant jet of tricuspid regurgitation (TRPG), adding the estimated right atrial pressure from the inferior vena cava (IVC). The tricuspid annulus plane systolic excursion was assessed using M-mode on the tricuspid annulus and expressed as the longitudinal systolic function of shortening the right ventricle.

### ESE

Following comprehensive transthoracic echocardiography at rest, patients underwent a symptom-limited graded exercise test in a semi-supine position on a bicycle ergometer table tilted to 20° as previously described (14). After maintaining an initial workload of 10 W for 3 min, the workload was increased by 10 W every 3 min. A single-lead electrocardiogram was continuously monitored, and blood pressure was measured at rest and every 1 min during exercise. Patients were excluded if they had the following abnormalities: (i) occurrence of angina, dizziness, or syncope; (ii) a decrease in systolic blood pressure below baseline; and (iii) complex arrhythmia during exercise.

### Endpoint

Follow-up data were collected from the medical records. The primary endpoint of the present study was the occurrence time of the first composite endpoint, defined as cardiovascular death, aortic valve replacement due to AS-related symptoms (syncope, dyspnea, and angina), and hospitalization for heart failure.

### Statistical analysis

Continuous variables are expressed as mean values with standard deviations, or median values with interquartile ranges (IQR). Categorical variables are expressed as numbers and percentages. Student’s t-test was used to analyze continuous variables with normal distributions, and the Mann–Whitney U-test was used to analyze continuous variables with non-normal distributions. Categorical variables were compared using Fisher’s exact or chi-square tests, as appropriate.

Statistical significance was set at p<0.05. The cumulative probability of event-free survival was estimated using the Kaplan–Meier method and compared between groups using a log-rank test. The optimal cutoff value of E/e’ during exercise was determined based on the receiver operating characteristics curve and highest Youden index. Univariable and multivariable Cox proportional hazard models were used to calculate hazard ratios and 95% confidence intervals for clinical outcomes. Data analyses were performed using JMP 16 (SAS Institute Japan, Inc., Tokyo, Japan) and R statistical software (version 4.2.2; R Foundation for Statistical Computing, Vienna, Austria).

## Results

Twenty-one patients were excluded due to the following reasons: (i) abnormal exercise response (n=4; atrial fibrillation tachycardia, advanced atrioventricular block during exercise, ventricular tachycardia, and ST change) and (ii) early intervention (n=17; transcatheter aortic valve replacement [TAVR] or surgical aortic valve replacement [SAVR] within 90 days after ESE). The remaining 122 patients, who were managed conservatively, were included in this study (Figure 1).

**Figure 1.**
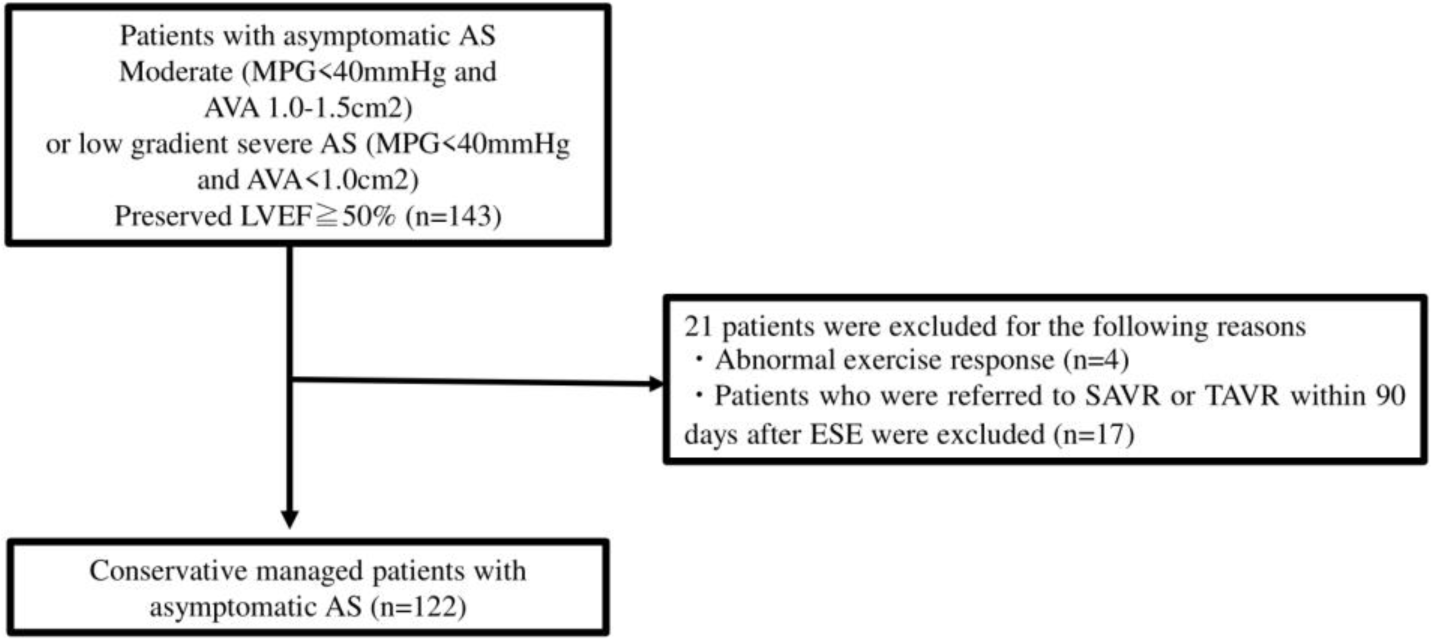
Flow chart showing study participant distribution AVA, aortic valve area; MPG, mean pressure gradient; LVEF, left ventricular ejection fraction; AS, aortic stenosis; ESE, exercise stress echocardiography; SAVR, surgical aortic valve replacement; TAVR, transcatheter aortic valve replacement

### Baseline characteristics

Table 1 shows the baseline characteristics of the study cohort. Fifty-nine (48.3%) patients were male, and the median age was 76.5 (IQR: 71.0-80.3) years. No significant differences in sex, age, comorbidities, or medications were observed between patients with moderate AS and those with low-gradient severe AS. However, patients with low-gradient severe AS had significantly smaller BSAs.

**Table 1.** Baseline clinical characteristics of the population.

| Variables | All<br>(n=122) | Moderate AS<br>(n=59) | LG severe AS<br>(n=63) | p-value |
| --- | --- | --- | --- | --- |
| Sex, male | 59 (48.3) | 33 (55.9) | 26 (41.3) | 0.105 |
| Age, years | 76.5 (71.0–80.3) | 76.0 (72.0–81.0) | 77.0 (70.0–80.0) | 0.912 |
| BSA, m<sup>2</sup> | 1.55 (1.42–1.71) | 1.60 (1.50–1.80) | 1.49 (1.39–1.63) | <0.001 |
| Hypertension | 80 (65.6) | 41 (69.5) | 39 (61.9) | 0.378 |
| DM | 24 (19.7) | 9 (15.3) | 15 (23.8) | 0.235 |
| DL | 52 (42.6) | 27 (45.8) | 25 (39.7) | 0.497 |
| CAD | 24 (19.7) | 12 (20.3) | 12 (19.1) | 0.858 |
| Hemodialysis | 5 (4.1) | 2 (3.4) | 3 (4.8) | 0.703 |
| AF | 29 (23.8) | 16 (27.1) | 13 (20.6) | 0.401 |
| Medication |  |  |  |  |
| β-blocker | 24 (19.7) | 15 (25.4) | 9 (14.3) | 0.122 |
| ACE inhibitor/ARB | 48 (39.3) | 24 (40.7) | 24 (38.1) | 0.770 |
| MRA | 4 (3.3) | 2 (3.4) | 2 (3.2) | 0.947 |
| Diuretics | 10 (8.2) | 6 (10.2) | 4 (6.4) | 0.442 |
| Bicuspid | 12 (9.9) | 6 (10.2) | 6 (9.5) | 0.905 |

Data are mean ±SD, median (IQR), or n (%).
LG, low-gradient; BSA, body surface area; DM, diabetes mellitus; DL, dyslipidemia; CAD, coronary artery disease; AF, atrial fibrillation; ACEi, angiotensin-converting enzyme inhibitor; ARB, angiotensin II receptor; MRA, mineralocorticoid receptor antagonist

### Resting and exercise echocardiography

The ESE data are summarized in Table 2 and 3. No significant differences in systolic blood pressure or HR were observed between the two groups at rest. However, patients with low-gradient severe AS had significantly lower AVA and AVA indices, higher MPG, and smaller left ventricular end-diastolic volume and left ventricular end-systolic volume than those with moderate AS. The LVEF was similar in both groups. SV and CO were significantly smaller in patients with low-gradient severe AS, possibly because of the LV size. The two groups had no significant differences in diastolic function, right ventricular function, or SPAP. Systolic blood pressure during exercise was comparable between the two groups. However, patients with low-gradient severe AS had significantly higher HR than those with moderate AS.

**Table 2.** Comparison of resting echocardiographic data between patients with moderate and low gradient severe AS.

| Variables | All<br>(n=122) | Moderate AS<br>(n=59) | LG severe AS<br>(n=63) | p-value |
| --- | --- | --- | --- | --- |
| SBP, mmHg | 140.1 (±21.2) | 141.5 (±20.9) | 138.9 (±21.5) | 0.507 |
| HR, beats/min | 69.0 (62.0–77.9) | 66.0 (62.0–73.0) | 71.0 (62.0–82.0) | 0.051 |
| AVA, cm<sup>2</sup> | 1.01 (±0.23) | 1.20 (±0.14) | 0.83 (±0.12) | <0.001 |
| AVA index, cm<sup>2</sup>/m<sup>2</sup> | 0.64 (±0.15) | 0.74 (±0.13) | 0.55 (±0.09) | <0.001 |
| MPG, mmHg | 20.5 (15.6–26.4) | 16.2 (13.4–22.1) | 24.4 (20.0–29.4) | <0.001 |
| LVDd, mm | 43.6 (±5.6) | 44.6 (±6.0) | 42.7 (±5.0) | 0.051 |
| LVDs, mm | 26.0 (24.0–29.3) | 27.0 (25.0–31.0) | 26.0 (24.0–29.0) | 0.058 |
| IVSd, mm | 9.0 (8.8–10.0) | 9.0 (8.0–10.0) | 9.0 (9.0–10.0) | 0.420 |
| PWd, mm | 9.0 (8.0–10.0) | 9.0 (8.0–10.3) | 9.0 (8.0–10.0) | 0.672 |
| LV mass index, g/m<sup>2</sup> | 81.8 (73.1–98.3) | 80.3 (71.1–97.3) | 82.1 (73.9–98.5) | 0.869 |
| SV, mL | 65.3 (58.0–74.1) | 69.0 (61.0–81.1) | 62.7 (54.0–72.0) | <0.001 |
| SVi, mL/m<sup>2</sup> | 42.5 (36.5–47.3) | 43.4 (38.8–47.3) | 41.5 (34.7–47.1) | 0.084 |
| Low-flow, (%) | 22 (18.0) | 6 (10.2) | 16 (25.4) | 0.026 |
| CO, L/min | 4.6 (3.9–5.3) | 4.9 (4.1–5.7) | 4.2 (3.7–5.1) | 0.027 |
| LVEDV, mL | 81.0 (67.6–99.0) | 85.0 (73.0–103.9) | 77.7 (67.0–94.0) | 0.053 |
| LVESV, mL | 26.7 (22.0–34.0) | 30.0 (22.5–39.5) | 25.0 (21.0–31.4) | 0.035 |
| LVEF, % | 66.1 (±6.0) | 65.3 (±6.7) | 66.7 (±5.2) | 0.208 |
| LAD, mm | 38.0 (33.0–42.0) | 38.0 (36.0–43.0) | 37.0 (32.0–42.0) | 0.295 |
| LAV index, mL/m<sup>2</sup> | 37.5 (30.0–45.8) | 37.4 (31.6–45.8) | 37.6 (28.7–45.8) | 0.877 |
| E, cm/sec | 73.0 (59.5–95.0) | 74.0 (58.0–94.0) | 69.0 (60.0–98.0) | 0.890 |
| e', cm/sec | 5.1 (4.3–6.1) | 5.3 (4.5–6.1) | 4.8 (4.1–6.0) | 0.171 |
| E/e' | 11.8 (9.5–16.6) | 13.4 (11.0–19.5) | 14.8 (11.3–19.8) | 0.573 |
| E/A | 0.74 (0.63–0.86) | 0.78 (0.66–0.89) | 0.73 (0.61–0.85) | 0.715 |
| Abnormal (%) | 78 (84.8) | 36 (85.7) | 42 (84.0) |  |
| Pseudonormal (%) | 14 (15.2) | 6 (14.3) | 8 (16.0) |  |
| ZVa, mmHg/ml/m<sup>2</sup> | 3.78 (3.30–4.44) | 3.68 (3.00–4.18) | 4.12 (3.48–4.50) | 0.010 |
| TRPG, mmHg | 23.0 (19.9–27.0) | 22.3 (18.9–27.5) | 23.5 (20.3–27.0) | 0.828 |
| SPAP, mmHg | 26.0 (22.9–30.1) | 25.3 (21.9–30.5) | 26.5 (23.3–30.0) | 0.838 |
| TAPSE, mm | 19.8 (16.9–22.1) | 19.9 (17.7–23.0) | 19.3 (16.7–21.5) | 0.484 |

Data are mean ± SD, median (IQR), or n (%).
AS, aortic stenosis; SBP, systolic blood pressure; HR, heart rate; AVA, aortic valve area; MPG, mean pressure gradient; LVDd, left ventricular diastolic dimension; LVDs, left ventricular systolic dimension; IVSd, interventricular septum dimension; PWd, left ventricular posterior wall dimension; LV, left ventricular; SV, stroke volume; SVi, stroke volume index; CO, cardiac output; LVEDV, left ventricular end-diastolic volume; LVESV, left ventricular end-systolic volume; LVEF, left ventricular ejection fraction; LAD, left atrial dimension; LAV, left atrial volume; ZVa, valvulo-arterial impedance; TRPRG, regurgitant jet of tricuspid regurgitation; SPAP, systolic pulmonary artery pressure; TAPSE, tricuspid annular plane systolic excursion

**Table 3.** Comparison of exercise echocardiographic data between the patients with moderate and low gradient severe AS.

| Variables | All<br>(n=122) | Moderate AS<br>(n=59) | LG severe AS<br>(n=63) | p-value |
| --- | --- | --- | --- | --- |
| Exercise intensity, METs | 3.5 (3.0–4.4) | 3.5 (2.9–4.3) | 3.7 (3.1–4.6) | 0.357 |
| Peak workload, W | 40 (30–57.5) | 40 (30–60) | 40 (30–50) | 0.200 |
| SBP, mmHg | 175.2 (±30.5) | 172.7 (±28.1) | 177.4 (±32.5) | 0.409 |
| HR, beats/min | 110.3 (±21.2) | 105.6 (±20.8) | 114.4 (±20.9) | 0.024 |
| AVA, cm<sup>2</sup> | 1.04 (0.92–1.25) | 1.24 (1.14–1.41) | 0.92 (0.79–1.02) | <0.001 |
| AVA index, cm<sup>2</sup>/m<sup>2</sup> | 0.66 (0.58–0.80) | 0.77 (0.64–0.92) | 0.59 (0.49–0.68) | <0.001 |
| MPG, mmHg | 26.4 (18.8–35.2) | 20.0 (16.6–27.8) | 31.6 (25.3–38.5) | <0.001 |
| SV, mL | 71.0 (61.3–83.2) | 74.2 (68.9–88.9) | 64.6 (55.4–78.0) | <0.001 |
| SVi, mL/m<sup>2</sup> | 46.3 (±10.6) | 49.0 (±10.9) | 43.8 (±9.8) | 0.006 |
| CO, L/min | 7.8 (±1.9) | 8.2 (±1.8) | 7.5 (±1.8) | 0.042 |
| LVEDV, mL | 86.0 (73.0–107.5) | 90.0 (74.0–109.7) | 81.4 (66.6–104.4) | 0.079 |
| LVESV, mL | 24.0 (18.0–30.9) | 25.0 (19.7–32.9) | 22.7 (17.0–28.9) | 0.330 |
| LVEF, % | 73.0 (68.5–76.0) | 73.0 (67.0–76.0) | 73.6 (69.0–75.9) | 0.635 |
| E, cm/sec | 127.0 (107.8–151.0) | 125.0 (103.0–149.0) | 131.0 (112.0–161.0) | 0.133 |
| e', cm/sec | 8.1 (6.9–10.3) | 8.0 (6.7–9.1) | 8.3 (6.9–12.5) | 0.011 |
| E/e' | 15.3 (11.8–18.8) | 14.9 (12.7–19.3) | 15.3 (10.3–18.0) | 0.816 |
| E/A | 1.04 (0.85–1.23) | 1.05 (0.83–1.24) | 0.97 (0.85–1.22) | 0.841 |
| Abnormal relaxation (%) | 46 (50.6) | 18 (43.9) | 28 (56.0) |  |
| pseudonormal (%) | 45 (49.5) | 23 (56.1) | 22 (44.0) |  |
| ZVa, mmHg/ml/m<sup>2</sup> | 4.48 (3.71–5.27) | 3.96 (3.29–4.91) | 4.95 (4.09–5.71) | <0.001 |
| TRPG, mmHg | 45.4 (±11.6) | 44.1 (±12.7) | 46.5 (±10.5) | 0.262 |
| SPAP, mmHg | 55.4 (±11.6) | 54.1 (±12.7) | 56.5 (±10.5) | 0.262 |
| Exercise-induced PH, (%) | 45 (38.5) | 22 (39.3) | 23 (37.7) | 0.861 |
| TAPSE, mm | 24.0 (±5.5) | 24.4 (±5.6) | 23.6 (±5.3) | 0.421 |
| SPAP/CO slope | 9.05 (6.40–14.46) | 7.76 (5.83–13.05) | 9.74 (7.18–15.42) | 0.049 |
| SPAP/TAPSE slope | 4.79 (2.75–9.91) | 4.85 (2.30–11.48) | 4.76 (3.15–8.36) | 0.471 |

Data are mean ± SD, median (IQR), or n (%).
AS, aortic stenosis; MET, metabolic equivalent; SBP, systolic blood pressure; HR, heart rate; AVA, aortic valve area; MPG, mean pressure gradient; LV, left ventricular; SV, stroke volume; SVi, stroke volume index; CO, cardiac output; LVEDV, left ventricular end-diastolic volume; LVESV, left ventricular end-systolic volume; LVEF, left ventricular ejection fraction; ZVa, valvulo-arterial impedance; TRPRG, regurgitant jet of tricuspid regurgitation; SPAP, systolic pulmonary artery pressure; TAPSE, tricuspid annular plane systolic excursion

AS severity parameters during exercise, including AVA and AVA index, were significantly lower, and the MPG was higher in patients with low-gradient severe AS, consistent with the resting data. The LV size during exercise was not significantly different between the two groups; however, patients with low-gradient severe AS had significantly smaller SV and CO during exercise.

### Comparisons between outcomes in low-gradient severe AS versus moderate AS

During a median follow-up period of 989 (IQR: 578–1571) days, the composite endpoint was occurred in 64 patients (cardiovascular death, n=2; hospitalization for heart failure, n=2; and AVR, n=58). Kaplan–Meier analysis revealed a significantly lower event-free survival rate in patients with low-gradient severe AS than in those with moderate AS (log-rank test, p<0.001; Figure 2). In multivariable Cox regression analysis revealed that low-gradient severe AS was independently associated with the event risk (hazard ratio=2.386, p=0.009 and p=0.002; Table 4).

**Figure 2.**
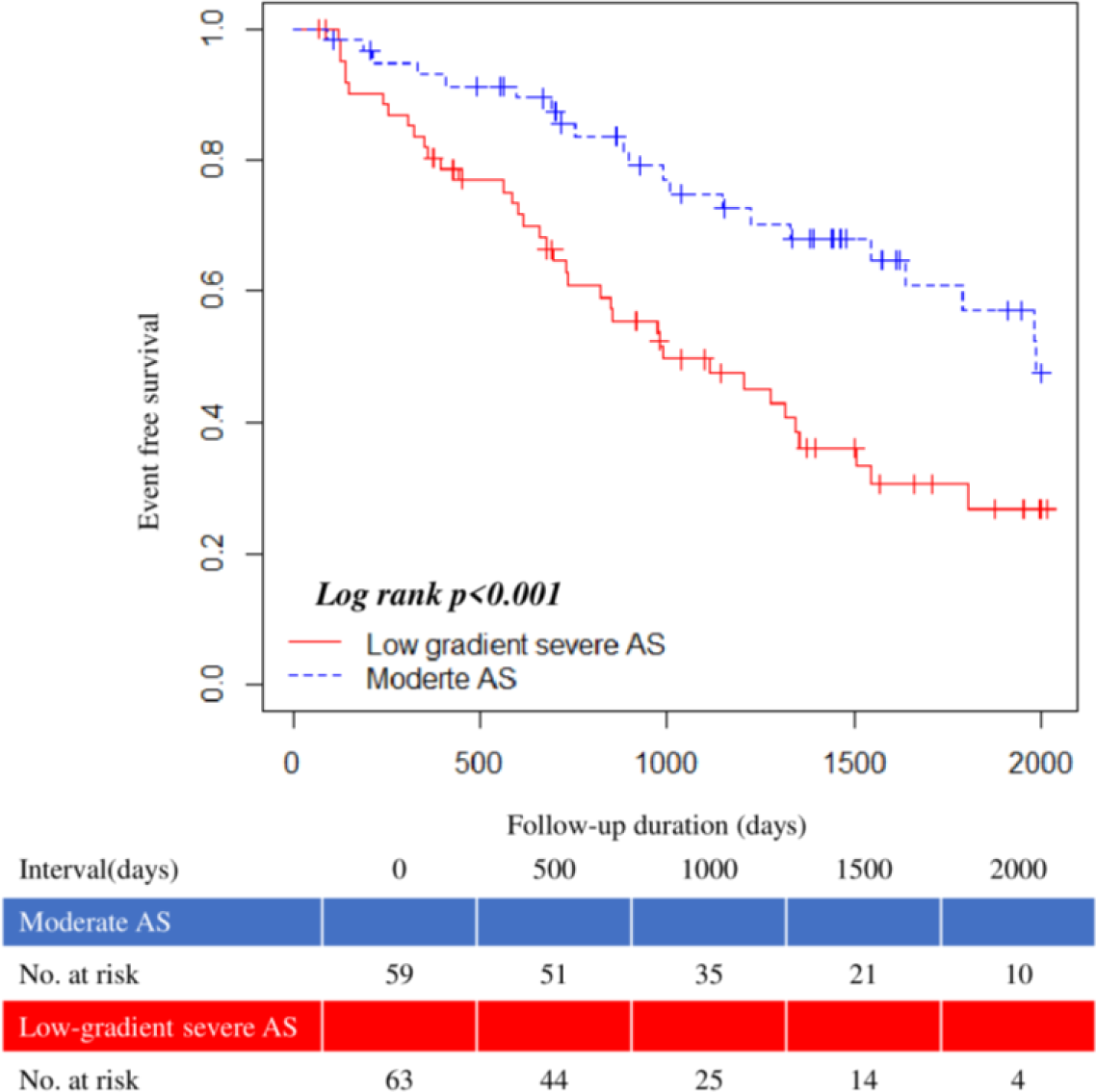
Event-free survival curve of moderate AS vs. low-gradient severe AS AS, aortic stenosis

**Table 4.**
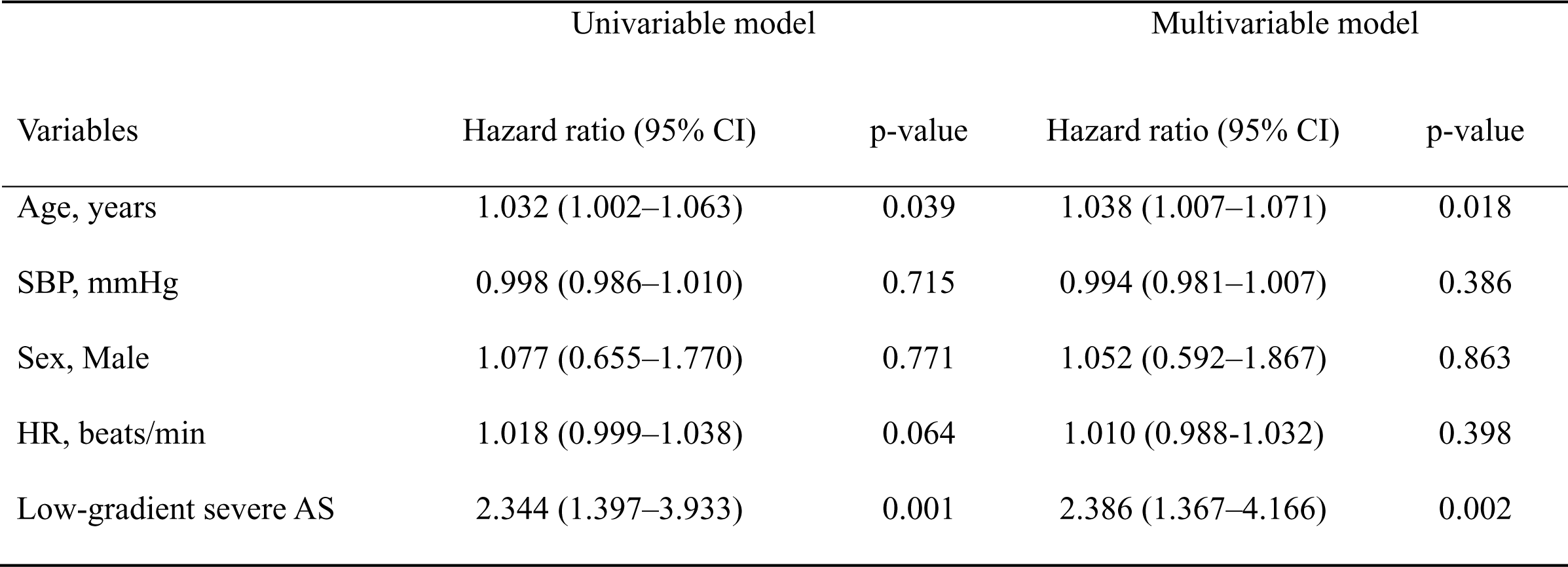
Univariable and multivariable Cox regression analyses for predicting aortic valve-related events in patients with moderate and low-gradient severe AS.

### Univariable and multivariable Cox regression analyses in continuous format

The prognostic factors in patients with low-gradient severe AS were investigated. The mitral E/e’ index during exercise was significantly associated with an increased risk of adverse events in the univariable analysis, using the continuous variable format, in patients with low-gradient severe AS (all p<0.01; Table 5). Contrastingly, the SPAP at rest and during exercise was not associated with the event risk.

**Table 5.**
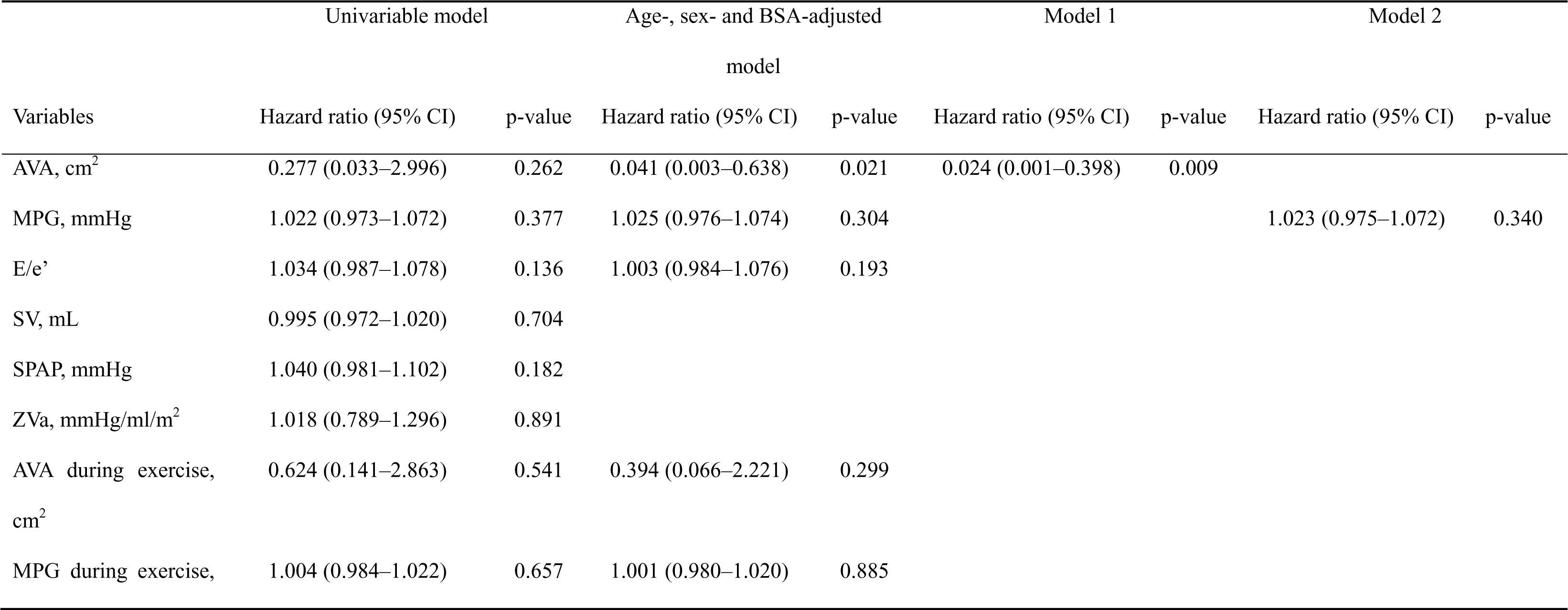
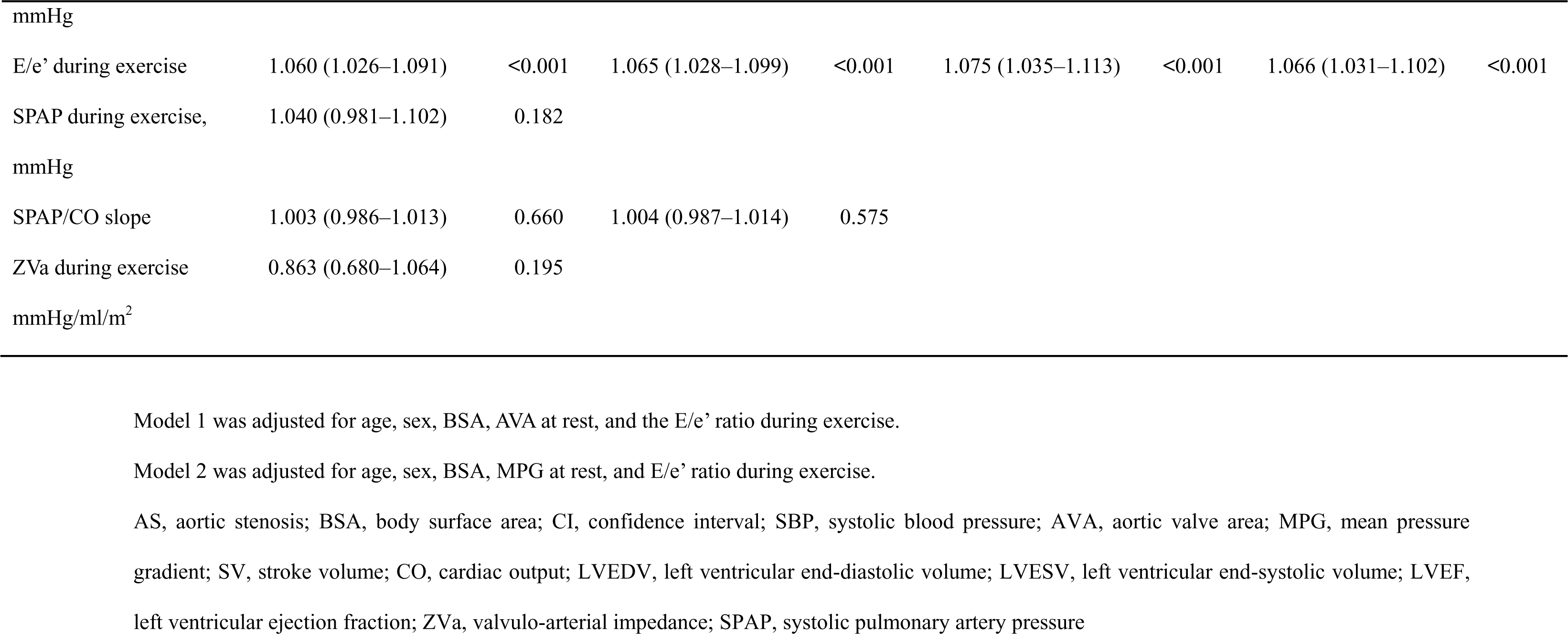
Univariable and multivariable Cox regression analyses for predicting aortic valve-related events in patients with low-gradient severe AS.

| Variables | Univariable model |  | Age-, sex- and BSA-adjusted model |  | Model 1 |  | Model 2 |  |
| --- | --- | --- | --- | --- | --- | --- | --- | --- |
|  | Hazard ratio (95% CI) | p-value | Hazard ratio (95% CI) | p-value | Hazard ratio (95% CI) | p-value | Hazard ratio (95% CI) | p-value |
| AVA, cm<sup>2</sup> | 0.277 (0.033–2.996) | 0.262 | 0.041 (0.003–0.638) | 0.021 | 0.024 (0.001–0.398) | 0.009 |  |  |
| MPG, mmHg | 1.022 (0.973–1.072) | 0.377 | 1.025 (0.976–1.074) | 0.304 |  |  | 1.023 (0.975–1.072) | 0.340 |
| E/e' | 1.034 (0.987–1.078) | 0.136 | 1.003 (0.984–1.076) | 0.193 |  |  |  |  |
| SV, mL | 0.995 (0.972–1.020) | 0.704 |  |  |  |  |  |  |
| SPAP, mmHg | 1.040 (0.981–1.102) | 0.182 |  |  |  |  |  |  |
| ZVa, mmHg/ml/m<sup>2</sup> | 1.018 (0.789–1.296) | 0.891 |  |  |  |  |  |  |
| AVA during exercise, cm<sup>2</sup> | 0.624 (0.141–2.863) | 0.541 | 0.394 (0.066–2.221) | 0.299 |  |  |  |  |
| MPG during exercise, | 1.004 (0.984–1.022) | 0.657 | 1.001 (0.980–1.020) | 0.885 |  |  |  |  |
| mmHg |  |  |  |  |  |  |  |  |
| E/e' during exercise | 1.060 (1.026–1.091) | <0.001 | 1.065 (1.028–1.099) | <0.001 | 1.075 (1.035–1.113) | <0.001 | 1.066 (1.031–1.102) | <0.001 |
| SPAP during exercise, | 1.040 (0.981–1.102) | 0.182 |  |  |  |  |  |  |
| mmHg |  |  |  |  |  |  |  |  |
| SPAP/CO slope | 1.003 (0.986–1.013) | 0.660 | 1.004 (0.987–1.014) | 0.575 |  |  |  |  |
| ZVa during exercise | 0.863 (0.680–1.064) | 0.195 |  |  |  |  |  |  |
| mmHg/ml/m<sup>2</sup> |  |  |  |  |  |  |  |  |

Model 1 was adjusted for age, sex, BSA, AVA at rest, and the E/e' ratio during exercise.
Model 2 was adjusted for age, sex, BSA, MPG at rest, and E/e' ratio during exercise.
AS, aortic stenosis; BSA, body surface area; CI, confidence interval; SBP, systolic blood pressure; AVA, aortic valve area; MPG, mean pressure gradient; SV, stroke volume; CO, cardiac output; LVEDV, left ventricular end-diastolic volume; LVESV, left ventricular end-systolic volume; LVEF, left ventricular ejection fraction; ZVa, valvulo-arterial impedance; SPAP, systolic pulmonary artery pressure

After adjusting for age, sex, and BSA, the AVA at rest and mitral E/e’ during exercise were significantly associated with an increased risk of adverse events. The variables for the multivariable Cox regression analysis were selected (Model 1: age, sex, BSA, AVA at rest, and mitral E/e’ index during exercise; Model 2: age, sex, BSA, MPG at rest, and mitral E/e’ index during exercise). Multivariable Cox regression analysis revealed that the AVA at rest and mitral E/e’ index during exercise were independently associated with the event risk (hazard ratio=0.024 and 1.075, p=0.009 and p<0.001, respectively; Table 5).

### Outcomes associated with the mitral E/e’ index during exercise in patients with low-gradient severe AS

The cutoff value for the mitral E/e’ index during exercise was set at 15.4 (ROC curve, p=0.007; AUC, 0.749; sensitivity, 65.9%; specificity, 86.3%). The patients in this study were divided into two groups based on their mitral E/e’ during exercise as follows: low E/e’ (<15.4, n=33) and high E/e’ (≥15.4, n=30) groups. Table 6 compares the clinical characteristics of patients with low-gradient severe AS between the low and high E/e’ groups. No significant differences in sex, age, BSA, and medications were observed between patients in the low and high E/e’ groups. The high E/e’ group had a higher prevalence of diabetes mellitus and coronary artery disease than the low E/e’ group.

**Table 6.** Comparison of clinical characteristics between patients with low-gradient severe AS in the low and high E/e’ groups.

| Variables | Low E/e'<br>(n=33) | High E/e'<br>(n=30) | p-value |
| --- | --- | --- | --- |
| Sex, male | 13 (39.4) | 13 (43.3) | 0.751 |
| Age, years | 77.0 (69.5–82.0) | 77.5 (72.5–80.0) | 0.673 |
| BSA, m<sup>2</sup> | 1.48 (1.39–1.61) | 1.51 (1.38–1.70) | 0.693 |
| Hypertension | 20 (60.6) | 19 (63.3) | 0.824 |
| DM | 4 (12.1) | 11 (36.7) | 0.021 |
| DL | 11 (33.3) | 14 (46.7) | 0.280 |
| CAD | 2 (6.1) | 10 (33.3) | 0.005 |
| Hemodialysis | 5 (15.2) | 8 (26.7) | 0.258 |
| AF | 5 (15.2) | 8 (26.7) | 0.259 |
| Medication | 4 (12.1) | 5 (16.7) | 0.607 |
| β-blocker | 15 (45.5) | 9 (30.0) | 0.205 |
| ACE inhibitor/ARB | 0 (0.0) | 2 (6.7) | 0.081 |
| MRA | 2 (6.7) | 2 (6.1) | 0.922 |
| Diuretics | 13 (39.4) | 13 (43.3) | 0.751 |
| Bicuspid | 77.0 (69.5–82.0) | 77.5 (72.5–80.0) | 0.673 |

Data are mean ±SD, median (IQR), or n (%).
AS, aortic stenosis; BSA, body surface area; DM, diabetes mellitus; DL, dyslipidemia; CAD, coronary artery disease; AF, atrial fibrillation; ACE, angiotensin converting enzyme; ARB, angiotensin receptor blocker; MRA, mineralocorticoid receptor antagonist

Table 7 summarizes the ESE data. The high E/e’ group had a significantly larger left atrial size, lower LVEF at rest, and lower AVA and AVA index during exercise than the low E/e’ group. Kaplan–Meier analysis indicated that the high E/e’ group had a significantly lower event-free survival rate than the low E/e’ group (log-rank, p=0.009; Figure 3). Prognostic stratification according to the mitral E/e’ index during exercise was possible in patients with low-gradient severe AS. Kaplan–Meier analysis stratified according to reduced (≤35 mL/m^2^, n=47) and preserved (>35 mL/m^2^, n=16) SVi showed no statistically significant difference in the rate of event-free survival (log-rank, p=0.860; Figure 4).

**Figure 3.**
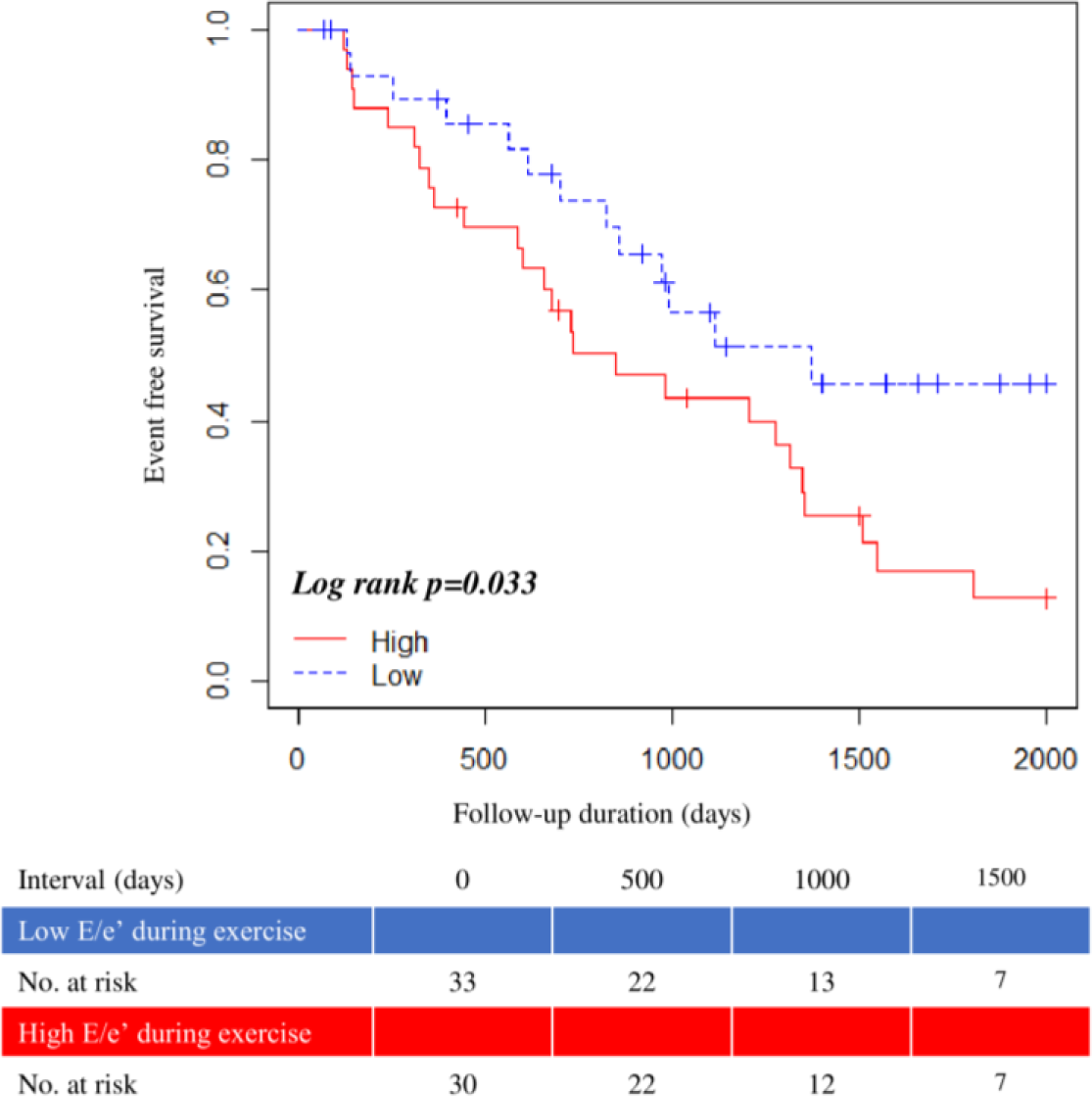
Event-free survival curve according to the mitral E/e’ in patients with low-gradient severe AS AS, aortic stenosis

**Figure 4.**
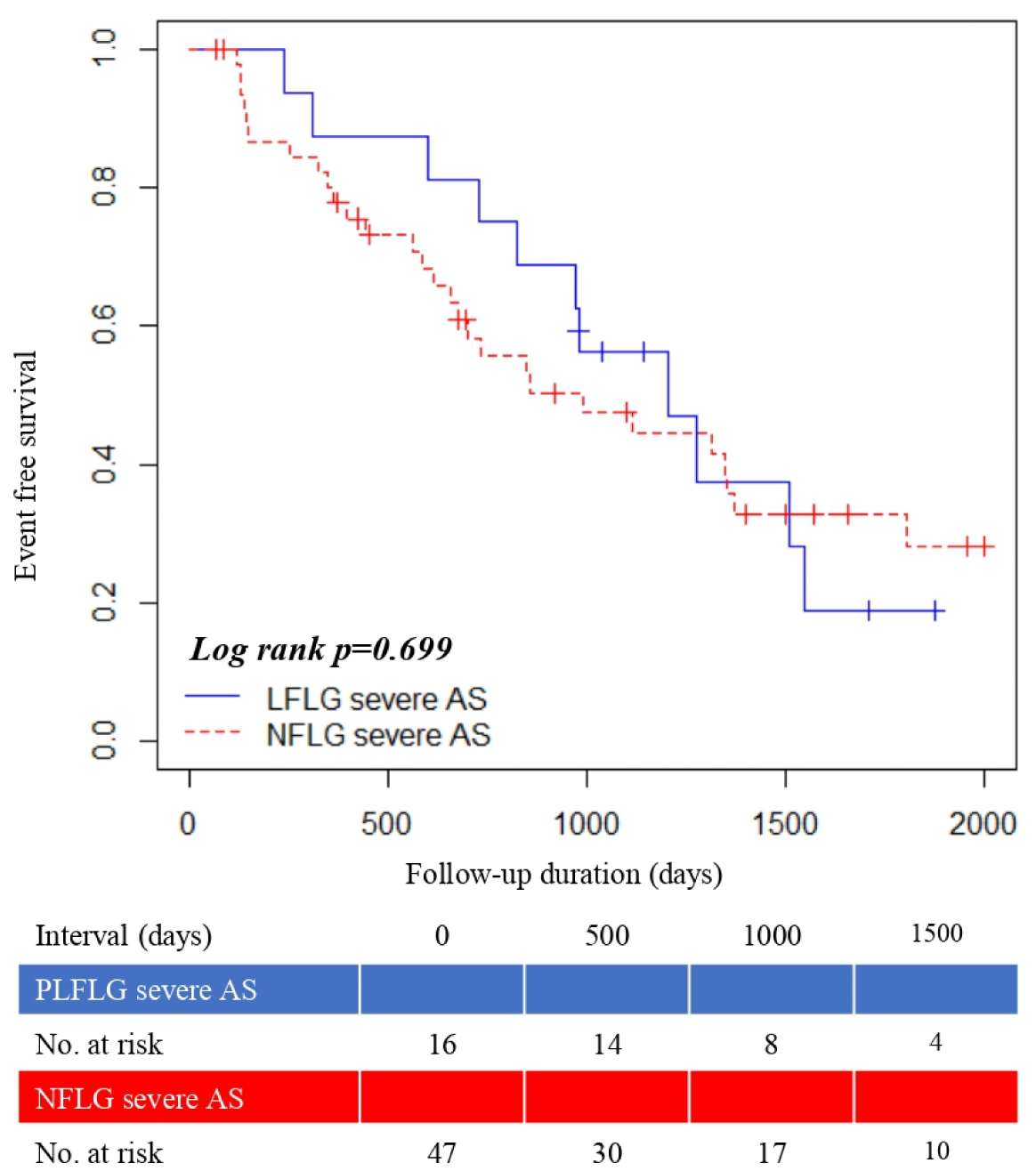
Event-free survival curve of reduced vs. preserved stroke volume index in patients AS, aortic stenosis; PLFLG, paradoxical low-flow low-gradient; NFLG, normal-flow low-gradient

**Table 7.** Comparison of resting and exercise echocardiographic findings between the low and high E/e’ during exercise in patients with low-gradient severe AS.

| Variables | Low E/e'<br>(n=33) | High E/e'<br>(n=30) | p-value |
| --- | --- | --- | --- |
| Exercise intensity, METs | 3.5 (3.1–4.9) | 3.7 (3.1–4.4) | 0.566 |
| Peak workload, M | 40.0 (30.0–55.0) | 46.5 (37.5–50.0) | 0.516 |
| **At rest** |  |  |  |
| SBP, mmHg | 142.9 (±23.0) | 134.6 (±19.3) | 0.133 |
| HR, beats/min | 71.0 (63.0–85.8) | 72.0 (58.0–82.0) | 0.600 |
| AVA, cm<sup>2</sup> | 0.84 (±0.11) | 0.81 (±0.13) | 0.262 |
| AVA index, cm<sup>2</sup>/m<sup>2</sup> | 0.56 (±0.09) | 0.53 (±0.09) | 0.172 |
| MPG, mmHg | 23.2 (19.3–29.8) | 24.6 (20.8–28.8) | 0.752 |
| LV mass index, g/m<sup>2</sup> | 80.2 (73.7–95.5) | 84.5 (74.7–107.9) | 0.417 |
| LVEDV, mL | 77.7 (67.3–94.8) | 77.0 (63.3–92.7) | 0.863 |
| LVESV, mL | 25.0 (21.1–30.8) | 25.3 (20.3–31.7) | 0.287 |
| LAV index, mL/m<sup>2</sup> | 34.3 (26.7–43.3) | 41.6 (31.1–52.6) | 0.011 |
| SV, mL | 62.7 (57.1–72.4) | 61.6 (49.8–71.3) | 0.444 |
| SVi, mL/m<sup>2</sup> | 41.7 (35.5–48.4) | 41.3 (32.6–45.7) | 0.325 |
| CO, L/min | 4.5 (3.8–5.3) | 4.1 (3.7–4.9) | 0.204 |
| LVEF, % | 68.1 (±4.6) | 65.2 (±5.4) | 0.028 |
| E, cm/sec | 65.0 (57.0–80.0) | 88.5 (63.0–114.8) | <0.001 |
| e', cm/sec | 4.8 (4.3–6.2) | 4.6 (3.9–5.9) | 0.261 |
| E/e' | 12.9 (9.6–15.4) | 19.6 (13.6–22.4) | <0.001 |
| E/A | 0.66 (0.58–0.85) | 0.75 (0.61–0.87) | 0.203 |
| Abnormal, % | 24 (85.7) | 18 (81.8) |  |
| Pseudonormal, % | 4 (14.3) | 4 (18.2) |  |
| SPAP, mmHg | 25.3 (23.1–28.4) | 27.9 (23.9–32.1) | 0.085 |
| TAPSE, mm | 19.8 (17.0–21.6) | 18.9 (16.0–21.4) | 0.243 |
| **During exercise** |  |  |  |
| SBP, mmHg | 186.7 (±29.9) | 166.9 (±32.7) | 0.016 |
| HR, beats/min | 117.0 (±23.1) | 111.5 (±18.0) | 0.294 |
| AVA, cm<sup>2</sup> | 0.96 (0.83–1.03) | 0.86 (0.69–0.99) | 0.050 |
| AVA index, cm<sup>2</sup>/m<sup>2</sup> | 0.63 (0.54–0.70) | 0.57 (0.48–0.64) | 0.025 |
| MPG, mmHg | 31.6 (24.8–38.4) | 31.9 (25.9–42.1) | 0.456 |
| SV, mL | 64.0 (57.5–80.1) | 65.7 (53.6–76.2) | 0.200 |
| SVi, mL/m<sup>2</sup> | 45.7 (±10.0) | 41.7 (±9.3) | 0.109 |
| CO, L/min | 7.9 (±1.7) | 7.1 (±1.9) | 0.079 |
| LVEF, % | 74.0 (69.9–76.8) | 72.4 (67.0–75.4) | 0.354 |
| E, cm/sec | 126.0 (104.0–141.5) | 140.0 (122.5–175.8) | 0.004 |
| e', cm/sec | 11.9 (7.8–14.2) | 7.3 (6.3–8.4) | <0.001 |
| E/e' | 10.7 (9.2–13.6) | 18.2 (16.6–23.0) | <0.001 |
| E/A | 0.93 (0.77–1.05) | 1.16 (0.92–1.50) | 0.004 |
| Abnormal, % | 21 (75.0) | 7 (31.8) |  |
| Pseudonormal, % | 7 (25.0) | 15 (68.2) |  |
| SPAP, mmHg | 55.8 (±10.7) | 57.3 (±10.5) | 0.599 |
| TAPSE, mm | 25.1 (21.4–27.0) | 24.3 (18.4–26.3) | 0.052 |

Data are mean ± SD, median (IQR), or n (%).
AS, aortic stenosis; MET, metabolic equivalent; SBP, systolic blood pressure; HR, heart rate; AVA, aortic valve area; MPG, mean pressure gradient; LV, left ventricular; LAV, left arterial volume; SV, stroke volume; SVi, stroke volume index; CO, cardiac output; LVEDV, left ventricular end-diastolic volume; LVESV, left ventricular end-systolic volume; LVEF, left ventricular ejection fraction; SPAP, systolic pulmonary artery pressure; TAPSE,
tricuspid annular plane systolic excursion

## Discussion

The main findings of this study are as follows: (i) asymptomatic patients with low-gradient severe AS had worse prognoses than those with moderate AS with only 40% event-free survival rate; and (ii) an increase in the mitral E/e’ index during exercise was effective for stratifying prognosis in asymptomatic patients with low-gradient severe AS.

### Prognosis of low-gradient severe AS

Previous studies have suggested that low-gradient severe AS might be caused by a reduced SV assessed using Doppler echocardiography, signifying an advanced stage of severe AS, compromised impaired ventricular function, and poor prognosis requiring early valve intervention (11,15). However, Nikolaus et al. (10) suggested that outcomes and progression rates in patients with low-gradient severe AS were comparable to those in patients with moderate AS. Contrastingly, the present study’s findings show worse prognoses in asymptomatic patients with low-gradient severe AS, potentially due to the inclusion of older and smaller patients with smaller LVs when compared to the study by Nikolaus et al. Several mechanisms could account for the low gradient in severe AS despite preserved LVEF. One possible mechanism could be reduced SV even with preserved LVEF, due to concentric LV hypertrophy or impaired longitudinal LV myocardial function.

Nonetheless, lower transvalvular pressure gradients may still occur in patients with small body sizes and LV dimensions. There could be several reasons for the low gradient, even though there was a severely stenotic AVA in the presence of preserved ejection fraction (EF). One possible reason is the outcome of reduced SV, even though EF is preserved, due to decreased ventricular size and/or impaired myocardial function (7,16). However, even when the SV is normal, patients with smaller body sizes and LV dimensions may still show a lower transvalvular pressure gradient (10).

### ESE for low gradient severe AS

Risk stratification of patients with asymptomatic AS remains controversial. ESE has emerged as an attractive risk stratification tool for distinguishing asymptomatic patients with AS. An increase in MPG >18–20 mmHg and increase in SPAP >60 mmHg during exercise have been suggested prognostic markers (17–19). However, the evidence remains limited, and Goublaire et al. reported that these parameters are ineffective for prognostic stratification (20). Furthermore, studies using stress echocardiography in patients with low-gradient severe AS are limited. Clavel et al. investigated the prognostic value of exercise and dobutamine stress echocardiography (DSE) in patients with severe PLFLG AS. They reported that prognostic stratification of patients with PLFLG severe AS was difficult using resting echocardiographic parameters, but projected AVA was effective for prognostic stratification (21). This study examined asymptomatic patients with ESE and symptomatic patients with DSE but did not discuss any parameters that reflect hemodynamics during exercise. It is the first study to examine ESE in patients with asymptomatic low-gradient severe AS, including echocardiographic index hemodynamics during exercise. Hemodynamic changes during exercise in patients with symptomatic, paradoxical, low-gradient severe AS have been previously studied using cardiopulmonary exercise testing combined with right heart catheterization and Doppler echocardiographic measurements. Peak oxygen consumption correlated inversely with the rate of pulmonary capillary wedge pressure (PCWP) increase. The ability to reduce vascular and valvular loads determines the effect of exercise on the PCWP. Baseline hemodynamic parameters did not predict the response to PCWP. Additionally, the hemodynamic responses did not show significant differences between patients with low flow (SVi ≤35 mL/m^2^) and those with normal flow (SVi >35 mL/m^2^) (22).

The current study investigated the relationship between prognosis and ESE parameters. MPG and EIPH, which were previously reported in asymptomatic AS, were difficult to use for prognostic stratification. However, the mitral E/e’ index during exercise was useful for prognostic stratification in patients with low-gradient severe AS. Changes in MPG and EIPH are affected by the SV and HR and may not be good prognostic indicators in low-gradient severe AS with reduced SV. In addition, it is difficult to assess whether a patient has EIPH because the right atrial pressure during exercise is difficult to estimate and is sometimes underestimated (23). The mitral E/e’ index correlates with LV stiffness and fibrosis (24,25) and is recognized as one of the indicators of LV diastolic function. In patients with heart failure with preserved ejection fraction (HFpEF), no increase in LV filling pressure was observed at rest; however, there were cases in which there was an increase in LV filling pressure with exercise. During exercise, in patients with HFpEF, impaired early diastolic relaxation, reduced increments in suction, and poor LV compliance result in insufficient increments in SV and CO, leading to increased LV filling pressure and higher PASP (26,27). The mitral E/e’ index correlates with invasively measured LVEDP during exercise (27). Therefore, the mitral E/e’ index during exercise is an important parameter in diagnosing HFpEF (28). A previous study showed that patients with symptomatic PLFLG with severe AS had a more advanced stage of diastolic dysfunction and significantly worse prognoses than those with asymptomatic PLFLG with severe AS (7,9). These results suggested a possible link between severe AS and HFpEF in some patients (29). Based on the results of the present study, patients with asymptomatic low-gradient severe AS who exhibited an increase in E/e’ during exercise, which is associated with HFpEF, were observed to have poorer prognoses.

### Limitations

This study had several limitations. First, this was a single-center, retrospective observational study, although long-term follow-up was available. Therefore, inherent bias cannot be excluded in this study type. Although the sample size was comparable to that of previous studies, it was relatively small, with a limited number of composite events. To date, no multicenter studies have been conducted on ESE in patients with low-gradient severe AS. Therefore, these results should be validated in multicenter prospective studies with larger populations. Second, the study population consisted of asymptomatic patients with moderate- or low-gradient severe AS who could undergo exercise testing. This study does not reflect low-gradient AS as a whole because it excluded patients who underwent SAVR or TAVR within 90 days of ESE and included asymptomatic patients. However, the results may reflect actual clinical practice since ESE is contraindicated in patients with symptomatic severe AS (30). Third, patients with abnormal exercise test findings and early intervention were excluded, and only those who were followed up with conservative treatment were included. Therefore, the study did not reflect the overall prognoses of patients with low-grade severe AS. Lastly, most of the events in this study were AVR, although cases with AVR within 90 days of ESE were excluded. The most of patients who underwent AVR were symptom onset and the indication was also determined by the heart-valve team. Waiting until heart failure hospitalization for AVR would be difficult in clinical practice, given the impact on patient outcomes.

## Conclusion

This study suggests that patients with low-gradient severe AS have worse prognoses than those with moderate AS. The mitral E/e’ ratio during exercise is a useful parameter for risk stratification of patients with low-gradient severe AS. LV diastolic function during exercise may play a crucial role in decision making for patients with low-gradient severe AS.

## Data Availability

Data will be shared on request to the corresponding author with permission of St. Marianna University Hospital.

## Acknowledgments

Department of Cardiology, St. Marianna University School of Medicine, Kawasaki, Japan.

## List of abbreviations

AS: aortic stenosis
SBP: systolic blood pressure
HR: heart rate
AVA: aortic valve area
MPG: mean pressure gradient
LVDd: left ventricular diastolic dimension
LVDs: left ventricular systolic dimension
IVSd: interventricular septum dimension
PWd: left ventricular posterior wall dimension
SV: stroke volume
SVi: stroke volume index
CO: cardiac output
LVESV: left ventricular end-systolic volume
LAD: left atrial dimension
LAV: left atrial volume
ZVa: valvulo-arterial impedance
TRPRG: regurgitant jet of tricuspid regurgitation
SPAP: systolic pulmonary artery pressure
TAPSE: tricuspid annular plane systolic excursion
PLFLG: paradoxical low-flow low-gradient
ESE: exercise stress echocardiography
SAVR: surgical aortic valve replacement
TAVR: transcatheter aortic valve replacement
DL: dyslipidemia
MRA: mineralocorticoid receptor antagonist
MET: metabolic equivalent

## References

1. Iung B, Baron G, Butchart EG et al. A prospective survey of patients with valvular heart disease in Europe: The Euro Heart Survey on Valvular Heart Disease. Eur Heart J 2003;24:1231–43.

2. Nkomo VT, Gardin JM, Skelton TN, Gottdiener JS, Scott CG, Enriquez-Sarano M. Burden of valvular heart diseases: a population-based study. Lancet 2006;368:1005–11.

3. Stewart BF, Siscovick D, Lind BK et al. Clinical factors associated with calcific aortic valve disease. Cardiovascular Health Study. J Am Coll Cardiol 1997;29:630–4.

4. Lindroos M, Kupari M, Heikkilä J, Tilvis R. Prevalence of aortic valve abnormalities in the elderly: an echocardiographic study of a random population sample. J Am Coll Cardiol 1993;21:1220–5.

5. Minners J, Allgeier M, Gohlke-Baerwolf C, Kienzle RP, Neumann FJ, Jander N. Inconsistencies of echocardiographic criteria for the grading of aortic valve stenosis. Eur Heart J 2008;29:1043–8.

6. Christensen KL, Ivarsen HR, Thuesen L, Kristensen B, Egeblad H. Aortic valve stenosis: fatal natural history despite normal left ventricular function and low invasive peak-to-peak pressure gradients. Cardiology 2004;102:147–51.

7. Hachicha Z, Dumesnil JG, Bogaty P, Pibarot P. Paradoxical low-flow, low-gradient severe aortic stenosis despite preserved ejection fraction is associated with higher afterload and reduced survival. Circulation 2007;115:2856–64.

8. Barasch E, Fan D, Chukwu EO et al. Severe isolated aortic stenosis with normal left ventricular systolic function and low transvalvular gradients: pathophysiologic and prognostic insights. J Heart Valve Dis 2008;17:81–8.

9. Dumesnil JG, Pibarot P, Carabello B. Paradoxical low flow and/or low gradient severe aortic stenosis despite preserved left ventricular ejection fraction: implications for diagnosis and treatment. Eur Heart J 2010;31:281–9.

10. Jander N, Minners J, Holme I et al. Outcome of patients with low-gradient “severe” aortic stenosis and preserved ejection fraction. Circulation 2011;123:887–95.

11. Otto CM, Nishimura RA, Bonow RO et al. 2020 ACC/AHA Guideline for the Management of Patients With Valvular Heart Disease: A Report of the American College of Cardiology/American Heart Association Joint Committee on Clinical Practice Guidelines. Circulation 2021;143:e72–e227.

12. Mitchell C, Rahko PS, Blauwet LA et al. Guidelines for Performing a Comprehensive Transthoracic Echocardiographic Examination in Adults: Recommendations from the American Society of Echocardiography. J Am Soc Echocardiogr 2019;32:1–64.

13. Briand M, Dumesnil JG, Kadem L et al. Reduced systemic arterial compliance impacts significantly on left ventricular afterload and function in aortic stenosis: implications for diagnosis and treatment. J Am Coll Cardiol 2005;46:291–8.

14. Hirasawa K, Izumo M, Suzuki K et al. Value of Transvalvular Flow Rate during Exercise in Asymptomatic Patients with Aortic Stenosis. J Am Soc Echocardiogr 2020;33:438–448.

15. Vahanian A, Beyersdorf F, Praz F et al. 2021 ESC/EACTS Guidelines for the management of valvular heart disease. Eur Heart J 2022;43:561–63213.

16. Cramariuc D, Rieck AE, Staal EM et al. Factors influencing left ventricular structure and stress-corrected systolic function in men and women with asymptomatic aortic valve stenosis (a SEAS Substudy). Am J Cardiol 2008;101:510–5.

17. Lancellotti P, Lebois F, Simon M, Tombeux C, Chauvel C, Pierard LA. Prognostic importance of quantitative exercise Doppler echocardiography in asymptomatic valvular aortic stenosis. Circulation 2005;112:I377–82.

18. Lancellotti P, Magne J, Donal E et al. Determinants and prognostic significance of exercise pulmonary hypertension in asymptomatic severe aortic stenosis. Circulation 2012;126:851–9.

19. Maréchaux S, Carpentier E, Six-Carpentier M et al. Impact of valvuloarterial impedance on left ventricular longitudinal deformation in patients with aortic valve stenosis and preserved ejection fraction. Arch Cardiovasc Dis 2010;103:227–35.

20. Goublaire C, Melissopoulou M, Lobo D et al. Prognostic Value of Exercise-Stress Echocardiography in Asymptomatic Patients With Aortic Valve Stenosis. JACC Cardiovasc Imaging 2018;11:787–795.

21. Clavel MA, Ennezat PV, Maréchaux S et al. Stress echocardiography to assess stenosis severity and predict outcome in patients with paradoxical low-flow, low-gradient aortic stenosis and preserved LVEF. JACC Cardiovasc Imaging 2013;6:175–83.

22. Pérez Del Villar C, Yotti R, Espinosa M et al. The Functional Significance of Paradoxical Low-Gradient Aortic Valve Stenosis: Hemodynamic Findings During Cardiopulmonary Exercise Testing. JACC Cardiovasc Imaging 2017;10:29–39.

23. Obokata M, Kane GC, Sorimachi H et al. Noninvasive evaluation of pulmonary artery pressure during exercise: the importance of right atrial hypertension. Eur Respir J 2020;55.

24. Kasner M, Westermann D, Steendijk P et al. Utility of Doppler echocardiography and tissue Doppler imaging in the estimation of diastolic function in heart failure with normal ejection fraction: a comparative Doppler-conductance catheterization study. Circulation 2007;116:637–47.

25. Kasner M, Westermann D, Lopez B et al. Diastolic tissue Doppler indexes correlate with the degree of collagen expression and cross-linking in heart failure and normal ejection fraction. J Am Coll Cardiol 2011;57:977–85.

26. Erdei T, Smiseth OA, Marino P, Fraser AG. A systematic review of diastolic stress tests in heart failure with preserved ejection fraction, with proposals from the EU-FP7 MEDIA study group. Eur J Heart Fail 2014;16:1345–61.

27. Burgess MI, Jenkins C, Sharman JE, Marwick TH. Diastolic stress echocardiography: hemodynamic validation and clinical significance of estimation of ventricular filling pressure with exercise. J Am Coll Cardiol 2006;47:1891–900.

28. Pieske B, Tschöpe C, de Boer RA et al. How to diagnose heart failure with preserved ejection fraction: the HFA-PEFF diagnostic algorithm: a consensus recommendation from the Heart Failure Association (HFA) of the European Society of Cardiology (ESC). Eur J Heart Fail 2020;22:391–412.

29. Namisaki H, Nagata Y, Seo Y et al. Symptomatic paradoxical low gradient severe aortic stenosis: A possible link to heart failure with preserved ejection fraction. J Cardiol 2019;73:536–543.

30. Atterhög JH, Jonsson B, Samuelsson R. Exercise testing: a prospective study of complication rates. Am Heart J 1979;98:572–9.

